# The plasmablast response to SARS-CoV-2 mRNA vaccination is dominated by non-neutralizing antibodies and targets both the NTD and the RBD

**DOI:** 10.1101/2021.03.07.21253098

**Authors:** Fatima Amanat, Mahima Thapa, Tinting Lei, Shaza M. Sayed Ahmed, Daniel C. Adelsberg, Juan Manuel Carreno, Shirin Strohmeier, Aaron J. Schmitz, Sarah Zafar, Julian Q Zhou, Willemijn Rijnink, Hala Alshammary, Nicholas Borcherding, Ana Gonzalez Reiche, Komal Srivastava, Emilia Mia Sordillo, Harm van Bakel, The Personalized Virology Initiative, Jackson S. Turner, Goran Bajic, Viviana Simon, Ali H. Ellebedy, Florian Krammer

## Abstract

In this study we profiled vaccine-induced polyclonal antibodies as well as plasmablast derived mAbs from individuals who received SARS-CoV-2 spike mRNA vaccine. Polyclonal antibody responses in vaccinees were robust and comparable to or exceeded those seen after natural infection. However, the ratio of binding to neutralizing antibodies after vaccination was greater than that after natural infection and, at the monoclonal level, we found that the majority of vaccine-induced antibodies did not have neutralizing activity. We also found a co-dominance of mAbs targeting the NTD and RBD of SARS-CoV-2 spike and an original antigenic-sin like backboost to seasonal human coronaviruses OC43 and HKU1. Neutralizing activity of NTD mAbs but not RBD mAbs against a clinical viral isolate carrying E484K as well as extensive changes in the NTD was abolished, suggesting that a proportion of vaccine induced RBD binding antibodies may provide substantial protection against viral variants carrying single E484K RBD mutations.

## Introduction

Understanding of the innate and adaptive immune responses to severe acute respiratory syndrome coronavirus 2 (SARS-CoV-2) has progressed rapidly since the beginning of the coronavirus disease 2019 (COVID-19) pandemic (Carvalho et al., 2021). Polyclonal antibody responses against the spike protein of the virus in serum, and to a lesser degree also at mucosal surfaces, have been well characterized with respect to their kinetics, binding capacity and functionality (Grandjean et al., 2020; Isho et al., 2020; Iyer et al., 2020; Ripperger et al., 2020; Seow et al., 2020; Wajnberg et al., 2020). Similarly, encouraging data have been published on both the plasmablast response and the memory B-cell response induced by SARS-CoV-2 infection (Dan et al., 2021; Gaebler et al., 2020; Guthmiller et al., 2021; Huang et al., 2021; Robbiani et al., 2020; Rodda et al., 2021; Wilson et al., 2020). The immune responses to SARS-CoV-2 vaccination, including to mRNA-based vaccines, are less well studied since these vaccines have only become available in the last months of 2020 (Baden et al., 2020; Polack et al., 2020). However, understanding vaccine-induced immunity is of high importance given the goal to achieve immunity for most people through vaccination, rather than as a consequence of infection.

The receptor binding domain (RBD) of the SARS-CoV-2 spike is an important target for serological and B-cell studies because it directly interacts with the cellular receptor angiotensin converting enzyme 2 (ACE2) mediating host cell entry (Letko et al., 2020; Wrapp et al., 2020). Antibodies binding to the RBD can potently block attachment of the virus to ACE2 and thereby neutralize the virus (Barnes et al., 2020). As a consequence, RBD-based vaccines are in development in addition to full length spike-based vaccines (Krammer, 2020). Analyses of the B-cell responses to the spike generally focus on the RBD and on cells sorted with RBD baits introducing an inherent bias by omitting non-RBD targets (Cao et al., 2020; Gaebler *et al*., 2020; Robbiani *et al*., 2020; Weisblum et al., 2020). This is also true for B cells and monoclonal antibodies (mAbs) isolated from vaccinated individuals (Wang et al., 2021). However, other epitopes within the spike protein, notably the N-terminal domain (NTD) but also S2, do harbor neutralizing epitopes (Chi et al., 2020; Liu et al., 2020; McCallum et al., 2021b; Song et al., 2020). In fact, the NTD is heavily mutated in the three most prominent variants of concern (VOCs, B.1.1.7, B.1.351 and P.1 (Davies et al., 2021; Faria et al., 2021; Tegally et al., 2020)). Here, we studied the unbiased plasmablast response to SARS-CoV-2 mRNA-based vaccination and report several new findings. First, we document that RBD and NTD co-dominate as B-cell targets on the viral spike protein, highlighting the importance of the NTD. We also report the first vaccine-induced NTD mAbs. In addition, we show that the majority of mAbs isolated are non-neutralizing, which is reflective of the higher binding to neutralization ratios found in serum after vaccination compared to natural infection. Finally, data from plasmablasts suggest that, at least, some of the vaccine-induced response is biased by pre-existing immunity to human β-coronaviruses.

## Results

### The polyclonal antibody response to mRNA vaccination exceeds titers seen in convalescent individuals but is characterized by a high ratio of non-neutralizing antibodies

In late 2020, six adult participants of an ongoing observational study received mRNA-based SARS-CoV-2 vaccines (**Suppl. Table 1**). Blood from these individuals (termed V1-V6) was collected at several time points including before vaccination (for 4/6), after the first vaccination and at several time points after the second vaccination. We examined their immune responses to recombinant spike protein and RBD in enzyme-linked immunosorbent assays (ELISA), in comparison to those of 30 COVID-19 survivors (**Figure 1A and 1B, Suppl. Table 1**). The sera from convalescent individuals were selected based on their anti-spike titers and grouped into three groups (low +: n=8; moderate ++: n=11; and high +++: n=11, based on the antibody titer measured in the Mount Sinai’s CLIA laboratory (Wajnberg *et al*., 2020)), in order to facilitate identifying different features that may track with the strength of the antibody response. Five out of six vaccinees produced anti-spike and anti-RBD responses that were, at the peak, markedly higher than responses observed even in the high titer convalescent group while one vaccinee (V4) produced titers comparable to the high titer group. Notably, the antibody response peaked one week after the second vaccine dose, followed by a decline in titers over the following weeks as expected from an antibody response to vaccination. Interestingly, anti-RBD antibody titers seemed to decline faster than anti-spike antibody titers, which appeared to be more stable over time. We also measured neutralizing antibody titers using authentic SARS-CoV-2 and found a similar trend with all vaccinees displaying high titers, even though V4 responded with delayed kinetics (**Figure 1C**). Importantly, although at the peak response, the vaccine group mounted neutralization titers that fell in the upper range for the high convalescent group, they did not exceed that group markedly. This finding prompted us to calculate the proportions of spike binding to neutralizing antibodies. For the convalescent group, we found that individuals with lower titers had a higher proportion of binding to neutralizing antibodies than high responding convalescent individuals (**Figure 1D**). When determined at the time of peak response, the vaccinees had the highest proportion of binding to neutralizing antibody titers, indicating an immune response focused on non-neutralizing antibodies or an induction of less potent neutralizing antibodies in general (or both). These proportions remained stable over time with the ratio of binding to neutralizing antibodies in vaccinated individuals being significantly higher than those observed for any of the three convalescent groups (p = 0.0004, 0.0002 and 0.0041 for the three groups respectively; **Suppl. Figure 1**). We also investigated the spike binding to RBD binding ratio and found no difference to convalescent individuals except a general trend towards proportionally less RBD binding over time in the vaccinees (**Suppl. Figure 1**).

**Figure 1:**
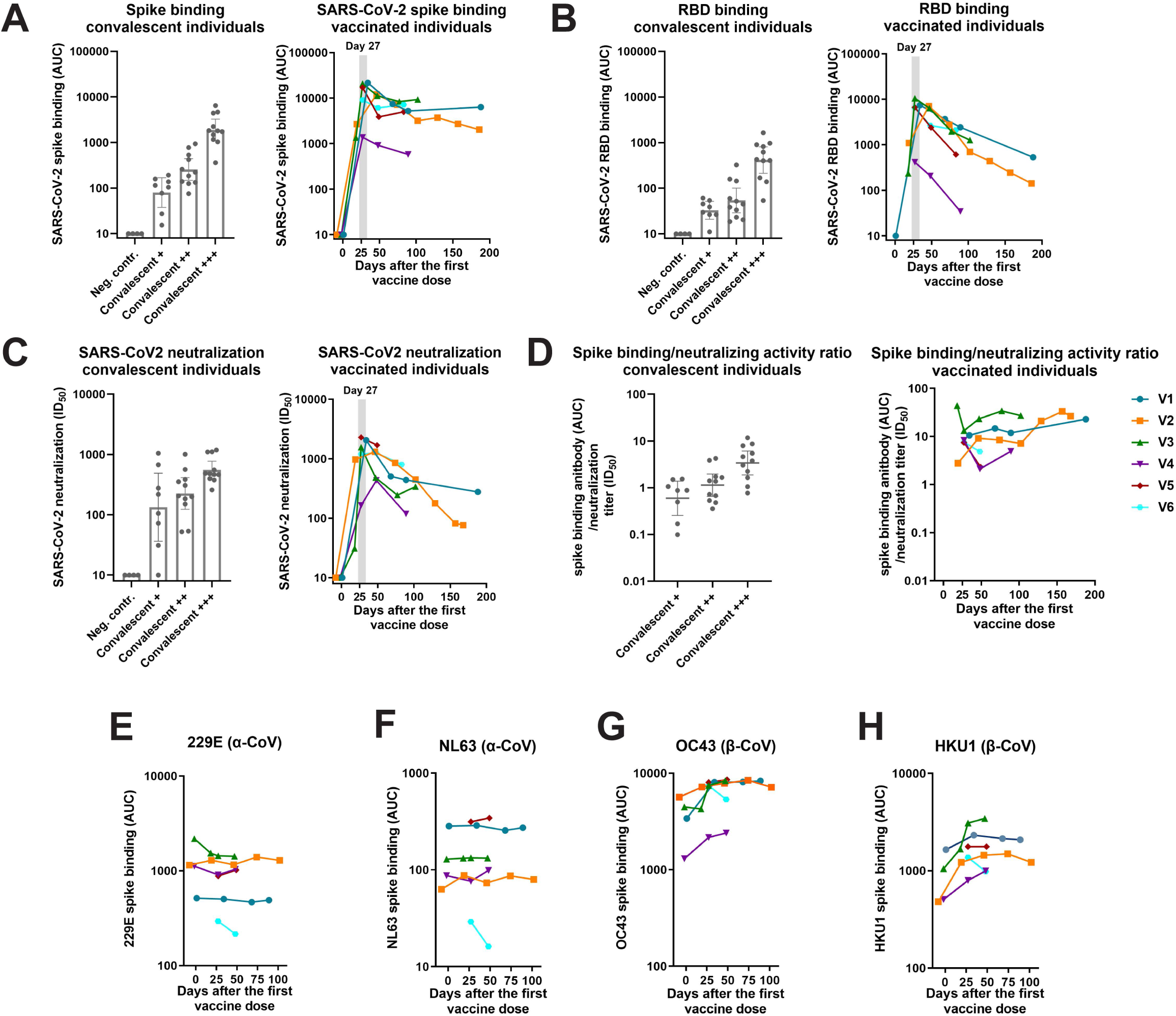
Antibody responses in individuals vaccinated with mRNA-based SARS-CoV-2 vaccines. Antibody responses of convalescent individuals and vaccinees to full length spike protein (**A**) and RBD (**B**) as measured by ELISA and neutralizing activity of the sera of the same individuals in a microneutralization assay against authentic SARS-CoV-2 (**C)**. Convalescent individuals were grouped based on their initial antibody response (measured in a CLIA laboratory) to spike protein into +, ++, and +++. **D** shows ratios between binding and neutralizing antibody levels in vaccinees and convalescent individuals. Higher ratios indicate a bias towards non-neutralizing antibodies. **E, F, G and H** show antibody responses against α-coronavirus 229E and NL63 and β-coronavirus OC43 and HKU1 spike proteins over time.

### mRNA vaccination induces a modest but measurable immune response to seasonal β-coronavirus spike proteins

It has been reported that SARS-CoV-2 infection induces an original antigenic sin-type immune response against human coronaviruses (hCoVs) to which the majority of the human population has pre-existing immunity (Aydillo et al., 2020; Song *et al*., 2020). Here, we explored whether this phenomenon is also induced by SARS-CoV-2 mRNA vaccination. Antibody titers in four vaccinees against spike protein from α-coronaviruses 229E and NL63 were detectable at the pre-vaccination time point, but did not increase substantially post-vaccination (**Figure 1E-F**; for V5 and V6 no pre-vaccination serum was available). However, titers against the spike proteins of β-coronaviruses OC43 and HKU1 increased substantially in these four vaccinees after vaccination (**Figure 1G-H)**. Thus, vaccination with mRNA SARS-CoV-2 spike also boosts immune responses against seasonal β-coronavirus spike proteins in a manner reminiscent of that reported for natural infection with SARS-CoV-2.

### Plasmablast response to SARS-CoV-2 mRNA vaccination targets both the RBD and the NTD

In order to characterize the B-cell response to vaccination in an unbiased manner, plasmablasts were single-cell sorted from blood specimens obtained from three individuals (V3, V5 and V6) one week after the booster immunization (**Suppl. Figure 2**). All mAbs were generated from single-cell sorted plasmablasts and probed for binding to recombinant SARS-CoV-2 spike protein. Twenty-one (40 mAbs were screened, with 28 being clonally unique, **Suppl. Table 2**) spike-reactive mAbs were isolated from V3, six (82 screened, 20 unique) from V5 and fifteen (84 screened, 24 unique) from V6 (**Figure 2A**). Using recombinant spike, RBD, NTD and S2 proteins, we mapped the domains to which these mAbs bind. Interestingly, only a minority of these antibodies recognized RBD (24% for V3, 47% for V6 and no RBD binders were identified for V5) (**Figure 2B and 2E**). A substantial number of the isolated mAbs bound to NTD including 14% for V3, 33% for V5 and 33% for V6 (**Figure 2C and 2E**). These data indicate that RBD and NTD are co-dominant in the context of mRNA-induced plasmablast response. The epitopes for the majority of the remaining spike binding mAbs, 52% for V3, 50% for V5 and 20% for V6, mapped to S2 (**Figure 2D and 2E**). Only three mAbs were not accounted for in terms of binding target (two for V3 and one for V5, **Figure 2E**).

**Figure 2.**
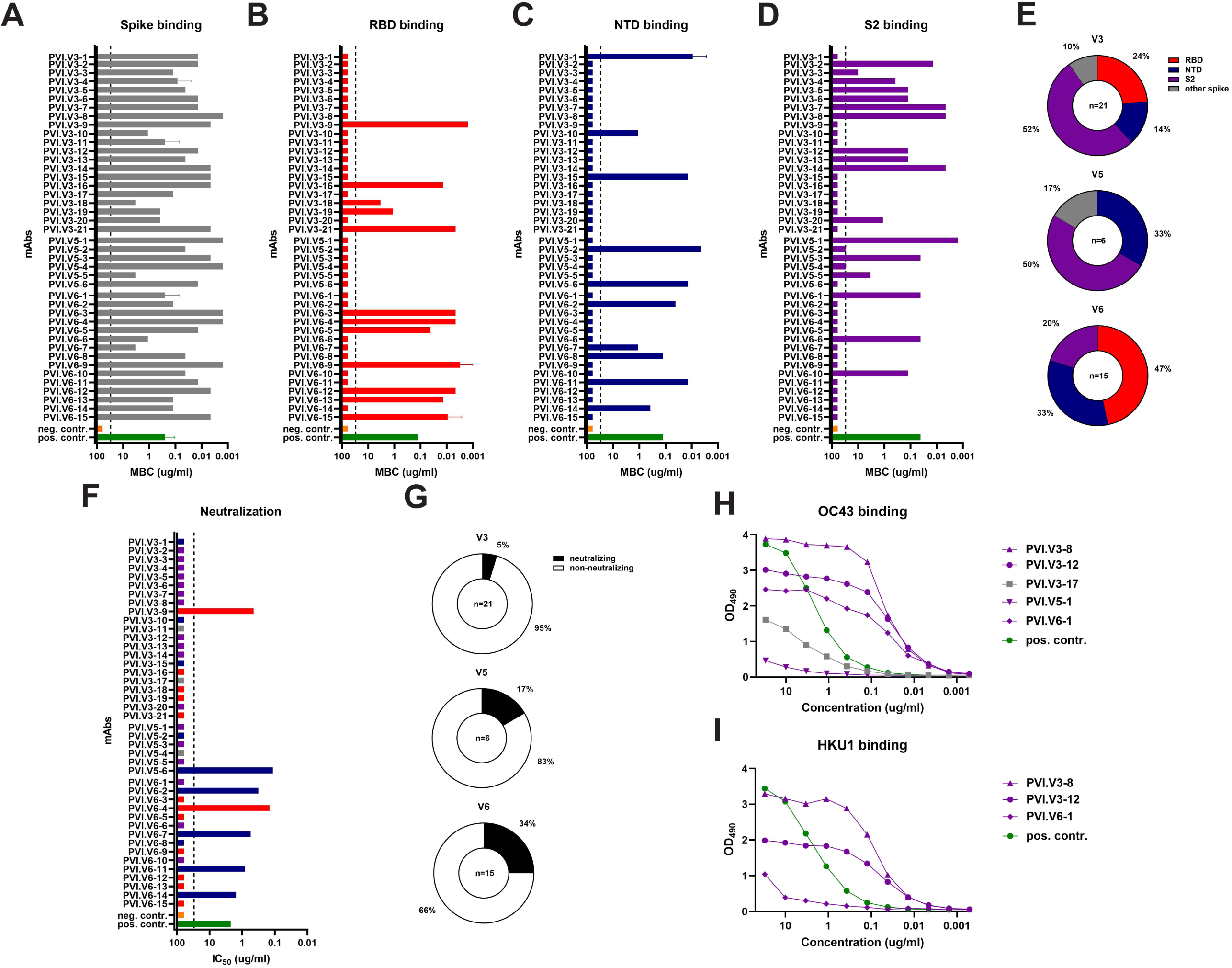
Characterization of mAbs derived from vaccine plasmablasts. Binding of plasmablasts derived from three vaccinees (V3, V5 and V6) against full length spike (**A**), RBD (**B**), NTD (**C**) and S2 (**D**). **E** shows the percentages of the respective antibodies per subject. **F** shows neutralizing activity of the mABs against authentic SARS-CoV-2 and the proportion of neutralizing antibodies per subject is shown in **G. H and I** show reactivity of mAbs to spike protein of human β-coronaviruses OC43 and HKU1. MBC = minimal binding concentration.

### The majority of isolated mAbs from SARS-CoV2 vaccinees are non-neutralizing

All antibodies were tested for neutralizing activity against the USA-WA1/2020 strain of SARS-CoV-2. Only a minority of the binding antibodies, even those targeting the RBD, showed neutralizing activity (**Figure 2F and 2G**). For V3, only one (an RBD binder) out of 21 mAbs (5%) displayed neutralizing activity (**Figure 2G**). For V5, a single NTD antibody neutralized authentic SARS-CoV-2 (17%) (**Figure 2G**). The highest frequency of neutralizing antibodies was found in V6 (34%) with one RBD neutralizer and four NTD neutralizers (**Figure 2G**). Interestingly, the highest neutralizing potency was found in mAb PVI.V5-6, an NTD binder followed by PVI.V6-4, an RBD binder.

We also tested all the antibodies for reactivity to the spike proteins of the four hCoVs 229E, NL63, HKU1 and OC43. No antibody binding to the spike proteins of α-coronaviruses 229E and NL63 was found but we identified five mAbs (including three from V3, one from V5 and one from V6) that bound, to varying degrees, to the spike of OC43, which, like SARS-CoV-2, is a β-coronavirus (**Figure 2H**). Three mAbs showed strong binding (PVI.V3-8, PVI.V3-12 and PVI.V6-1), while PVI.V3-17 showed an intermediate binding phenotype and PVI-V5-1 bound very weakly. Three of these mAbs also showed binding to the spike of HKU1, another β-coronavirus. Of these, PVI.V6-1 showed only very weak binding while PVI.3-8 and PVI.3-12 had low minimal binding concentrations (MBCs) indicating higher affinity (**Figure 2I**).

### The spike-reactive plasmablast response is dominated by IgG1+ cells and is comprised of a mixture of cells with low and high levels of somatic hypermutation (SHM)

Single-cell RNA sequencing (scRNAseq) was performed on bulk sorted plasmablasts from the three vaccinees (V3, V5, V6) to comprehensively examine the transcriptional profile, isotype distribution and somatic hypermutation (SHM) of vaccine-induced plasmablasts. We analyzed 4,584, 3,523 and 4,461 single cells from subjects V3, V5, and V6, respectively. We first verified the identity of sequenced cells as plasmablasts through the combined expression of B cell receptors (BCRs) (**Figure 3A**) and that of the canonical transcription as well as other factors essential for plasma cell differentiation, such as *PRDM1, XBP1* and *MZB1* (**Figure 3B**). To identify vaccine-responding B cell clones among the analyzed plasmablasts, we used scRNAseq to also analyze gene expression and V(D)J libraries from the sorted plasmablasts and clonally matched the BCR sequences to those from which spike-specific mAbs had been made. Using this method, we recovered 332, 7 and 1,384 BCR sequences from the scRNAseq data that are clonally related to the spike-binding mAbs derived from subjects V3, V5 and V6, respectively (**Figure 3C**). It is important to note here that we were not able to recover clonally related sequences for all of the mAbs that we cloned and expressed from each of the three vaccinees.

**Figure 3.**
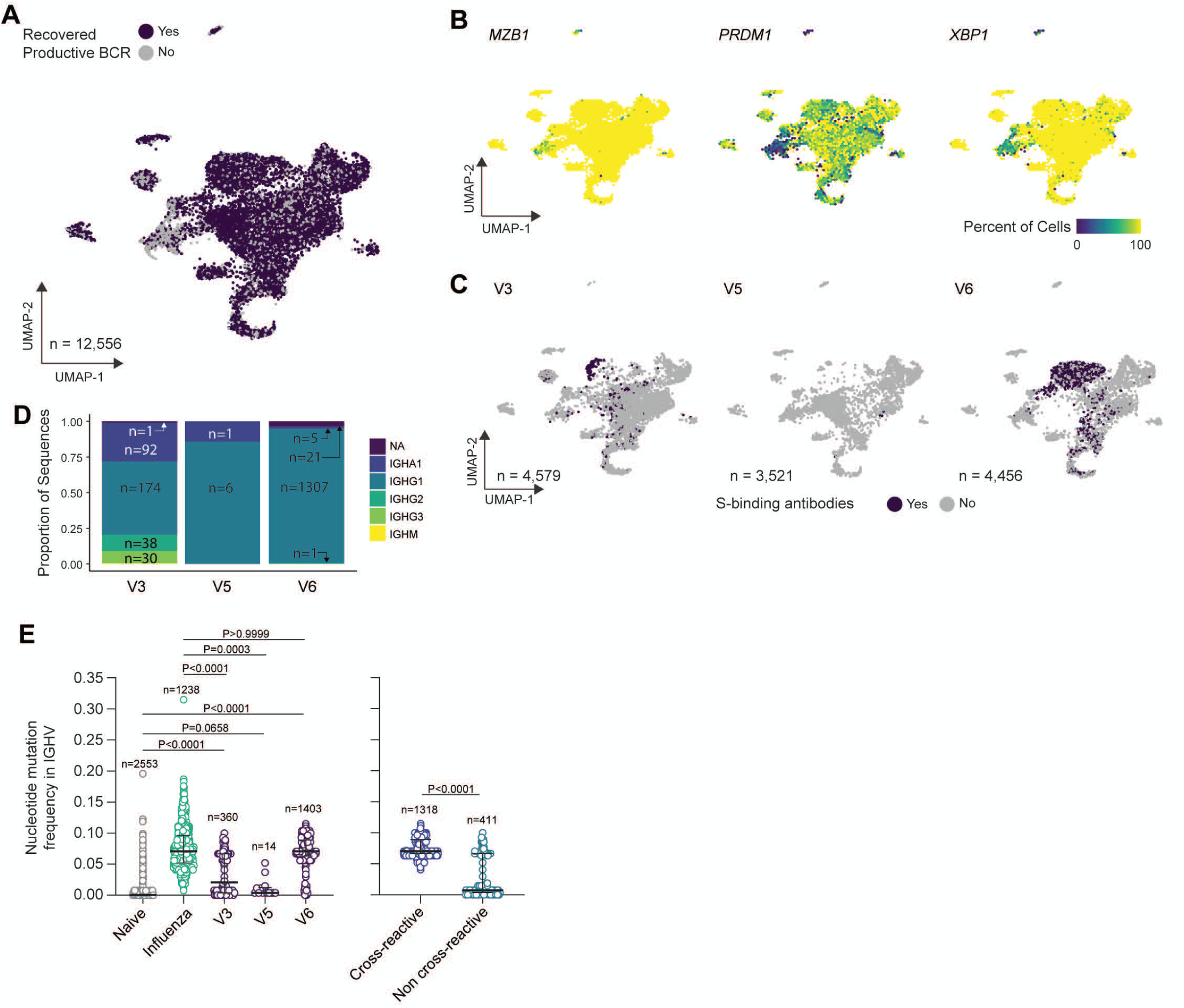
Characterization of bulk sorted plasmablasts via single-cell RNA sequencing. **(A)** Uniform manifold approximation and projection (UMAP) of scRNAseq from bulk plasmablast with recovered BCR sequences (purple) or unrecovered (grey). **(B)** UMAP overlay of percent of cellular population expressing *MZB1, PRDM1*, and *XPB1*. Hexbin equals 80 individual cells. (**C**) UMAP overlay of BCR sequences with confirmed spike binding activity. (**D**) Proportional composition of heavy chains genes in the spike binding sequences broken down by sample. **(E)** Comparison of nucleotide-level mutation frequency in immunoglobulin heavy chain variable (IGHV) genes between plasmablasts clonally related to spike binding mAbs from SARS-CoV-2 vaccinees, plasmablasts sorted from PBMCs one week after seasonal influenza vaccination and found in vaccine-responding B cell clones, and naïve B cells found in blood of an influenza vaccinee (left panel); and between plasmablasts from SARS-CoV-2 vaccinees found to be clonally related to spike-binding mAbs that were, respectively, cross-reactive and non-cross-reactive to human β-coronaviruses spike proteins (right panel).

We next examined the isotype and IgG subclass distribution among the recovered sequences. IgG1 was by far the most dominant isotype in the three vaccinees (**Figure 3D**). Finally, we assessed the level of somatic hypermutation (SHM) among the mAbs-related sequences from the three subjects. We used the SHM levels observed in human naïve B cells and seasonal influenza virus vaccination-induced plasmablasts that were previously published for comparison (Turner et al., 2020). Spike-reactive plasmablasts from V3 and V6 but not V5 had accumulated SHM at levels that are significantly greater than those observed with naïve B cells (**Figure 3E, left panel**). Strikingly, the SHM among V6 plasmablasts was equivalent to those observed after seasonal influenza virus vaccination (**Figure 3E, left panel**). We reasoned that the high level of SHM among spike-reactive plasmablasts may be derived from those targeting conserved epitopes that are shared with human β-coronaviruses. Indeed, we found that the SHM level among clones that are related to cross-reactive mAbs was significantly higher than their non-cross-reactive counterparts (**Figure 3E, right panel**).

### Competition of RBD binding neutralizing mAbs with ACE2 and affinity of variant RBDs for human ACE2

Two mAbs were identified as neutralizing and binding to RBD. We wanted, therefore, to test if they competed with ACE2 for RBD binding. Concentration-dependent competition was indeed observed for both mAbs demonstrating that inhibition of ACE2 binding is the mechanism of action of the two mAbs (**Figure 4**). Since we prepared RBD proteins of viral variants of concern for analysis of antibody binding (see below), we also wanted to assess the affinity of each variant RBD for human ACE2. Using biolayer interferometry (BLI), we measured association and dissociation rates of the N501Y RBD mutant (B.1.1.7 carries that mutation as its sole RBD mutation), Y453F, as found in mink isolates (Larsen et al., 2021), N439K, which is found in some European clades (Thomson et al., 2021), a combination of Y453F and N439K, E484K (part of B.1.351 and P.1) as well as for the B.1.351 and the P.1 RBDs for a recombinant version of human ACE2 (**Figure 4A, 4B and 4D**). Almost all of the single and double mutations in RBD tested increased affinity to human ACE2. Specifically, N501Y and Y453F combined with N439K increased affinity for human ACE2 by 5-fold (**Figure 4D, Suppl. Figure 3**). In contrast, E484K on its own decreased affinity by 4-fold. Of note, the B.1.351 RBD affinity for ACE2 was comparable to that of the wild-type RBD. These data were confirmed using an ELISA-based method which showed the same trends (**Suppl. Figure 4**).

**Figure 4.**
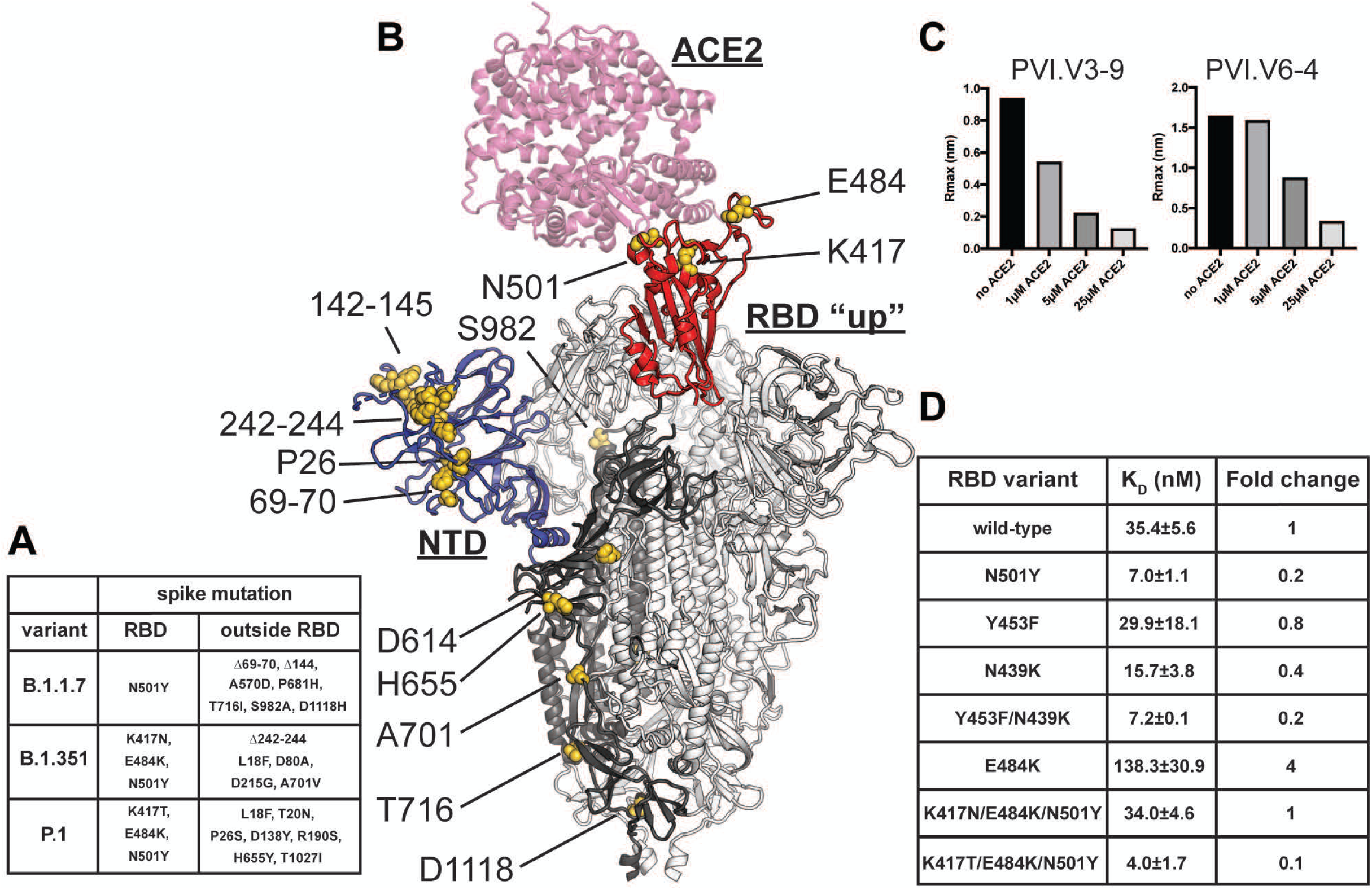
Mapping of the amino-acid substitutions and deletions onto the structure of the SARS-CoV-2 spike glycoprotein. **A** lists mutations of the three major variants of concern B.1.17, B.1.315 and P.1. **B** shows these mutations mapped onto the structure of the spike glycoprotein (model generated by superposition of PDB 6M0j and 7C2L (Chi *et al*., 2020; Lan et al., 2020)). One RBD in the up conformation (red) is bound with ACE2 receptor (pink). The NTD is colored blue and the various amino-acid substitutions are shown as yellow spheres. One spike protomer is shown in bold colors while the other two are colored white. **C** shows competition between ACE2 and neutralizing RBD targeting mAbs PVI.V3-9 and PVI.V6-4 for binding to RBD. **D** BLI-measured binding affinities of the RBD mutants to ACE2, as well as the calculated fold change, are shown in the table on the right.

### Binding profiles of polyclonal serum and mAbs to RBDs carrying mutations found in viral variants of concern

Next, we assessed binding of sera from vaccinated individuals, COVID-19 survivors and mAbs derived from plasmablasts to variant RBDs. Our panel of RBDs includes published mAb escape mutants, RBD mutants detected by the Mount Sinai Hospital’s Pathogen Surveillance Program in patients seeking care at the Mount Sinai Health System in NYC as well as mutations found in viral variants of interest and variants of concern (Baum et al., 2020; Greaney et al., 2021; Larsen *et al*., 2021; Thomson *et al*., 2021; Weisblum *et al*., 2020). Serum from convalescent individuals showed strong fluctuations depending on the viral variant (**Figure 5A**). In general, single mutants E406Q, E484K and F490K exerted the biggest impact on binding. However, complete loss of binding was rare and 2-4-fold reduction in binding was more common. Interestingly, almost all sera bound better to N501Y RBD (B.1.1.7) than to wild-type RBD (average 129% compared to wild type). Conversely, the B.1.351 RBD caused, on average, a 39% reduction in binding. The impact was slightly lower for the P.1 RBD (average 70% binding compared to wild-type). For sera from the six vaccinated individuals, however, the highest reduction seen was only two-fold for E406Q, N440K, E484K and F490K (**Figure 5B**). Of note, the vaccinees’ later samples (V1=d89, V2=d102, V3=d47, V4=d48, V5=49 and V6=48) were assayed to allow for some affinity maturation. The highest reduction observed for E484K, F484A, B.1.351 and P.1 were also approximately two-fold but this did not apply to all six vaccinees. Some vaccinees maintained binding levels against these RBDs at levels comparable to wild-type RBD.

**Figure 5.**
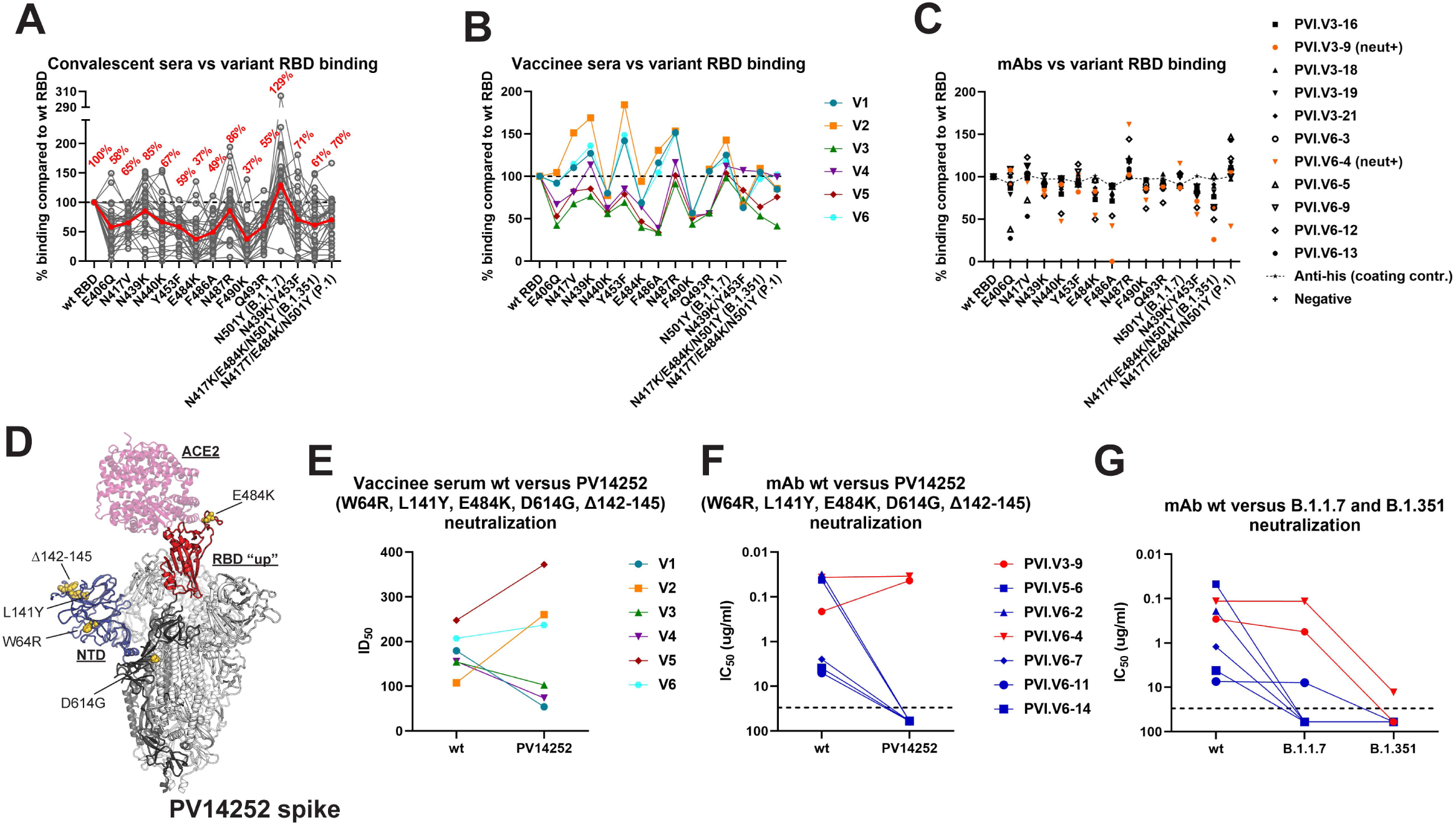
Binding and neutralization of SARS-CoV-2 variants. Binding of serum samples from convalescent individuals, vaccinees and vaccine derived mAbs to a panel of RBD mutants is shown in **A, B and C** respectively. The red line in A indicates the average reduction. Dotted lines in A and B indicate 100%, the line with smaller dots in C indicated reactivity of the anti-his coating control. For vaccinees late samples (V1=d89, V2=d102, V3=d47, V4=d48, V5=49 and V6=48) were assayed. **D** shows the spike mutations of virus isolate PVI14252 modelled on a co-crystal structure of the SARS-CoV-2 spike protein with ACE2 (model generated by superposition of PDB 6M0j and 7C2L (Chi *et al*., 2020; Lan *et al*., 2020)). **E and F** show the inhibitory effect of vaccine serum and vaccine derived neutralizing antibodies on both wild type SARS-CoV-2 and PV14252. **G** shows neutralizing activity of the plasmablast derived neutralizing antibodies aginst wild type, B.1.1.7 and B.1.351 virus isolates. Of note, these comparative assays were always performed side by side but sets are run by different operators and on a different Vero cell clone as the neutralization assays shown in Figure 2.

RBD binding mAbs were also tested for binding to the same variants. In general, mAbs maintained binding levels within 2-fold of the binding seen with the wild-type RBD with some exceptions. In fact, for most mAbs, no impact on binding was observed (**Figure 5C**) with the exception of PVI.V3-9, which lost binding to the RBD carrying F486A. Although there was a negative impact on binding of several mAbs to the B.1.351 variant, binding was almost unaffected by the mutations in the P.1 variant RBD. Only one mAb, PVI.V6-4, showed a drop in binding to P.1.

### Escape of an NTD and E484K mutant virus from polyclonal post-vaccination serum is negligible but NTD mutations significantly impact the neutralizing activity of NTD binding mAbs

Through the Mount Sinai Hospital’s Pathogen Surveillance Program, we had access to the SARS-CoV-2 isolate PV14252 (Clade 20C, Pango lineage B.1) that featured two mutations (W64R, L141Y) and one deletion (Δ142-145) in the NTD as well as the E484K mutation in the RBD (**Figure 5D**). To determine the susceptibility of this virus variant to neutralization by post-vaccination serum, we performed microneutralization assays. Wild-type SARS-CoV-2 and PV14252 were tested in parallel to ensure that the assay setup for both viruses allowed comparison. We found a relatively minor impact when testing polyclonal sera from vaccinees for neutralizing activity (**Figure 5E**). The activity of sera from V2, V5 and V6 slightly increased while the activity for V1, V3 and V4 decreased. Next, we tested the seven neutralizing mAbs that we isolated from plasmablasts. Consistent with their binding profiles in the variant RBD ELISA, the two RBD mAbs neutralized both viruses with comparable efficiency (**Figure 5F**). In fact, the activity of PVI.V3-9 increased slightly (**Figure 5F**). In stark contrast, all five anti-NTD antibodies completely lost neutralizing activity against PV14252 due to mutations present in the NTD of this viral isolate.

### B.1.1.7 and B.1.351 partially escape from plasmablast derived neutralizing antibodies

We also tested the neutralizing activity of the two RBD and the five NTD antibodies against the variants of concern B.1.1.7 and B.1.351 (**Figure 5G**). Both variants contain deletions as well as mutations in the NTD. In addition, B.1.1.7 carries the N501Y RBD mutation and B.1.351 carries N417K, E484K and N501Y mutations in the RBD (**Figure 4 A and B**). The two RBD binding antibodies lost no (PVI.V6-4) or little (PVI.V3-9) neutralizing activity against B.1.1.7. However, PVI.V3-9 lost all activity against B.1.351 and the remaining neutralizing activity of PVI.V6-4 was low (but measurable). All but one (PVI.V6-11)) NTD mAbs lost neutralizing activity against B.1.1.7 and all of them lost neutralizing activity against B.1.351 once more highlighting the importance of changes in the NTD on the antibody activity.

## Discussion

Our knowledge of B-cell responses to SARS-CoV-2 mRNA vaccination remains incomplete. We urgently need information about the nature of polyclonal vaccine-induced responses as well as unbiased, in depth analyses of plasmablast responses. Our data provide important new insights into these responses in comparison with immune responses to natural infection. Indeed, SARS-CoV-2 infection results in a very heterogeneous antibody response to the spike protein in terms of antibody quantity. In contrast, mRNA vaccination appears to induce a high antibody response of relatively homogenous titers. However, we also found that vaccinees generate more non-neutralizing antibodies than COVID-19 survivors resulting in a lower ratio of neutralizing to binding antibodies. These data were already apparent in the early phase clinical trials but remained unrecognized at the time (Walsh et al., 2020). Interestingly, low titer convalescent serum had the highest relative amount of neutralizing antibodies, whereas the proportion of binding antibodies was increased in sera with higher measured antibody titers. The majority of plasmablasts sampled after vaccination do, in fact, produce non-neutralizing antibodies. Two recent studies have performed a similarly unbiased plasmablast analysis for individuals naturally infected with SARS-CoV-2 (Cho et al., 2021; Huang *et al*., 2021). Of course, the antibody response after SARS-CoV-2 infection is not only targeting the spike protein but several other proteins expressed by the virus. When accounting for spike binding only, these studies report proportions of 44% and 25% neutralizing antibodies (Cho *et al*., 2021; Huang *et al*., 2021). While plasmablast analysis is in general not quantitative (e.g. one clone per clonotype is selected etc.) our analysis of post-vaccination plasmablasts found a lower number of neutralizing antibodies (17%).

Future studies are needed to reveal the role of non-neutralizing antibodies in SARS-CoV-2 immune protection. Indeed, antibody functions other than neutralization have been shown to correlate with protection (Bartsch et al., 2021; Gorman et al., 2021; Schäfer et al., 2021). The importance of absolute antibody titers and not ratios is underscored by the fact that post-vaccination neutralization titers were equal to or exceeded the titers found in the high responder convalescent group.

Of the four seasonal CoVs that are widely circulating in humans, β-coronaviruses OC43 and HKU1 have higher homology to SARS-CoV-2 spike. Vaccinated individuals mounted a response to spike proteins from OC43 and HKU1 but not to α-coronaviruses 229E and NL63. This phenomenon resembles the immune imprinting described in influenza virus immunology and has already been shown for natural infection with SARS-CoV-2 where a ‘backboost’ to β-coronaviruses was also found (Aydillo *et al*., 2020; Song *et al*., 2020). A few of the mAbs isolated in our study had, indeed, such a cross-reactive phenotype. It remains unclear whether these antibodies, which target mostly S2 epitopes, contribute to protection against SARS-CoV-2, OC43 or HKU1 infection. However, the cross-reactive epitopes of mAbs that do bind SARS-CoV-2, HKU1 and OC43 spikes could provide the basis for future pan-β-coronavirus vaccines. While it is likely the case that the B-cells producing these mAbs come from recall responses and were initially induced by human β-coronaviruses (which is supported by serology and of course the extensive SHM that the mAbs show), they could hypothetically also be *de novo* induced antibodies. While this is probably not the case, we cannot exclude this possibility with our current data.

Another interesting point we noted is the co-dominance of RBD and NTD. Previous analyses of B-cell responses to SARS-CoV-2 mRNA vaccination focused on cells baited by labeled RBD (Wang *et al*., 2021). We, in contrast, took an unbiased approach to sort and clone plasmablasts in an antigen-agnostic manner. We found similar levels of NTD and RBD binders with many mAbs binding to epitopes outside the RBD and the NTD. In one vaccinee not a single RBD binding mAbs was isolated with the caveats that the overall number of mAbs derived from that individual were low and their polyclonal serum antibody responses included RBD recognition. These data suggest that the NTD, which also harbors neutralizing epitopes, is - at least - as important as the RBD and warrants as much attention. In fact, five out of seven neutralizing antibodies isolated in this study bound to the NTD and only two targeted the RBD. Recent studies analyzing the plasmablast response after natural infection have found a similar co-dominance of RBD and NTD (Cho *et al*., 2021; Huang *et al*., 2021) with one study reporting 59 mAbs targeting the RBD, 64 targeting the NTD and 46 binding outside of RBD and NTD and the second study finding 10 RBD mAbs, 13 non-RBD S1 binding mAbs (strongly suggesting NTD binding) and 9 mAbs targeting S2. Further characterization of the mAbs obtained in our study showed a complete loss of neutralization against an authentic, replication-competent variant virus that harbored extensive changes in the NTD. All NTD mAbs also lost neutralizing activity against B.1.351 and all but one lost activity against B.1.1.7. These observations may explain why a reduction in neutralization against the viral variant of concern B.1.1.7 is seen in some studies despite the fact the N501Y substitution in the RBD of this variant does not significantly impact binding and neutralizing activity (Emary et al., 2021). The key role of NTD as target for antibodies has recently also been shown using memory B cell derived mAbs (McCallum et al., 2021a).

In addition, we assessed the impact of different RBD mutations on affinity towards human ACE2. Interestingly, N501Y increased the affinity by five-fold. This increase in receptor binding affinity may contribute to the higher infectivity of B.1.1.7, which carries this mutation in its RBD. In contrast, introduction of E484K reduced the affinity by 4-fold which may explain why virus variants carrying only the E484K mutation have rarely spread efficiently, although viruses carrying E484K have been detected since the fall of 2020 in a handful of patients receiving care at the Mount Sinai Health System and have also been reported in immunocompromised patients (Choi et al., 2020). It is tempting to speculate that the N501Y mutation enables the acquisition of E484K without a fitness loss. In fact, the B.1.351 RBD, which carries N501Y and E484K (as well as N417K) showed binding to hACE2 that was similar to wild-type RBD. Recently, B.1.1.7 variant strains carrying E484K, in addition to N501Y, have been isolated in the UK (PHE, 2021), providing evidence for the hypothesis that N501Y enables acquisition of mutations in the RBD that may be detrimental to receptor binding. However, recent expansion of B.1.526, a lineage also featuring E484K but without N501Y in New York City, suggests that this fitness loss may be overcome by other, yet uncharacterized, changes in the virus as well (Annavajhala et al., 2021; Lasek-Nesselquist et al., 2021). Interestingly, binding of convalescent sera to the N501Y RBD was also increased, suggesting that changes that increase affinity for the receptor may also increase affinity of a set of antibodies that may mimic the receptor.

We also noted that the two neutralizing antibodies against the RBD showed some reduced binding to a mutant RBD carrying the E484K mutation while having similar or even increased neutralizing potency against a variant virus carrying the E484K mutation as the only change in its RBD. The reduced affinity of the E484K variant RBD for hACE2 could render the virus more susceptible to RBD binding mAbs. Thus, an antibody binding to the RBD may just be more effective in interfering with a low affinity as compared to a high affinity RBD-hACE2 interaction. Increased affinity as an escape mechanism for viruses has been described in the past (Hensley et al., 2009; O’Donnell et al., 2012) and the converse mechanism could be at play here.

Whether or not the current vaccines will provide effective protection against circulating and emerging viral variants of concern is an important question which has gathered a lot of attention in early 2021. Our data indicate that reduction in binding to the E484K and B.1.351 variant RBDs was minor (often only 2-fold) compared to reported reduction in neutralization (which ranges from 6-8 fold to complete loss of neutralization (Cele et al., 2021; Wibmer et al., 2021; Wu et al., 2021)). Although not tested here, it is likely that the reduction in binding to full length spike is even lower, given the many epitopes on the spike other than NTD and RBD. The maintenance of binding to a large degree observed in this study suggests that viral variants will have a minor impact on serological assays which are currently in wide use for medical, scientific and public health reasons. Binding, non-neutralizing antibodies have also been shown to have a protective effect in many viral infections (Asthagiri Arunkumar et al., 2019; Dilillo et al., 2014; Saphire et al., 2018) and may be a factor in the substantial residual protection seen in the Johnson & Johnson and Novavax vaccine trials against B.1.351 in South Africa (Shinde et al., 2021). Production of non-neutralizing antibodies may also play a role in protection by mRNA vaccines after the first dose, as it is substantial and occurs during a time when neutralizing antibody titers are either very low or absent (Baden *et al*., 2020; Dagan et al., 2021; Polack *et al*., 2020). Finally, although some antibodies may lose neutralizing activity due to reduced affinity, they do still bind. Furthermore, B cells with these specificities potentially could undergo affinity maturation after exposure to a variant virus or a variant spike-containing vaccine, leading to high affinity antibodies to variant viruses of concern.

In summary, we demonstrate that the antibody responses to SARS-CoV-2 mRNA vaccination comprise a large proportion of non-neutralizing antibodies and are co-dominated by NTD and RBD antibodies. The NTD portion of the spike represents, thus, an important vaccine target. Since all viral variants of concern are heavily mutated in this region, these observations warrant further attention to optimize SARS-CoV-2 vaccines. Finally, broadly cross-reactive mAbs to β-coronavirus spike proteins are induced after vaccination, and suggest a potential development path for a pan-β-coronavirus vaccine.

## Data Availability

Data is available upon reasonable request from the corresponding authors.

## Acknowledgements

We would like to thank the study participants for their generosity and willingness to participate in longitudinal COVID19 research studies. None of this work would be possible without their contributions. We would like to thank Dr. Randy A. Albrecht for oversight of the conventional BSL3 biocontainment facility, which makes our work with live SARS-CoV-2 possible. We are also grateful for Mount Sinai’s leadership during the COVID-19 pandemic. We want to especially thank Drs. Peter Palese, Carlos Cordon-Cardo, Dennis Charney, David Reich and Kenneth Davis for their support. We would also like to thank Bassem Mohamed and Wooseob Kim for their help with preparing the scRNAseq libraries. This work was partially funded by the NIAID Collaborative Influenza Vaccine Innovation Centers (CIVIC) contract 75N93019C00051, NIAID Center of Excellence for Influenza Research and Surveillance (CEIRS, contract # HHSN272201400008C and HHSN272201400006C), NIAID grants U01AI141990 and U01AI150747, by the generous support of the JPB Foundation and the Open Philanthropy Project (research grant 2020-215611 (5384); and by anonymous donors. J.S.T. was supported by NIAID 5T32CA009547.

## Materials and methods

### Human subjects and specimen collection

The study protocols for the collection of clinical specimens from individuals with and without SARS-CoV-2 infection by the Personalized Virology Initiative were reviewed and approved by the Mount Sinai Hospital Institutional Review Board (IRB-16-16772; IRB-16-00791; IRB-20-03374). All participants provided written informed consent prior to collection of specimen and clinical information. All specimens were coded prior to processing and analysis. An overview of the characteristics of the vaccinees as well as the study participants with and without COVID-19 is provided in **Suppl. Table 1**. The vaccinees received two doses of the Pfizer mRNA vaccine.

Whole blood was collected via phlebotomy in serum separator tubes (SST) or ethylenediaminetetraacetic acid (EDTA) tubes. Serum was collected after centrifucation as per manufacturers’ instructions. Peripheral blood mononuclear cells (PBMCs) isolation was performed by density gradient centrifugation using SepMate tubes (Stemcell) according to manufacturers’ instructions. PBMCs were cryo-preserved and stored in liquid nitrogen until analysis.

### Recombinant proteins

All recombinant proteins were produced using Expi293F cells (Life Technologies). Receptor binding domain (RBD) and spike protein of SARS-CoV-2 (GenBank: MN908947.3) was cloned into a mammalian expression vector, pCAGGS as described earlier (Amanat et al., 2020b; Stadlbauer et al., 2020). RBD mutants were generated in the pCAGGS RBD construct by changing single residues using mutagenesis primers. All proteins were purified after transient transfections with each respective plasmid. Six-hundred million Expi293F cells were transfected using the ExpiFectamine 293 Transfection Kit and purified DNA. Supernatants were collected on day four post transfection, centrifuged at 4,000 g for 20 minutes and finally, the supernatant was filtered using a 0.22 um filter. Ni-NTA agarose (Qiagen) was used to purify the protein via gravity flow and proteins were eluted as previously described (Amanat *et al*., 2020b; Stadlbauer *et al*., 2020). The buffer was exchanged using Amicon centrifugal units (EMD Millipore) and all recombinant proteins were finally re-suspended in phosphate buffered saline (PBS). Proteins were also run on a sodium dodecyl sulphate (SDS) polyacrylamide gels (5–20% gradient; Bio-Rad) to check for purity (Amanat et al., 2018; Margine et al., 2013). Plasmids to express recombinant spike proteins of 229E, HKU1, NL63 and OC43 were generously provided by Dr. Barney Graham (Pallesen et al., 2017). NTD and S2 proteins were purchased from SinoBiologics.

### ELISA

Ninety-six well plates (Immulon 4 HBX; Thermo Scientific) were coated overnight at 4°C with recombinant proteins at a concentration of 2 ug/ml in PBS (Gibco; Life Technologies) and 50 uls/well. The next day, the coating solution was discarded. One hundred uls per well of 3% non-fat milk prepared in PBS (Life Technologies) containing 0.01% Tween-20 (TPBS; Fisher Scientific) was added to the plates to block the plates for 1 hour at room temperature (RT). All serum dilutions were prepared in 1% non-fat milk prepared in TPBS. All serum samples were diluted 3-fold starting at a dilution of 1:50. After the blocking step, serum dilutions were added to the respective plates for two hours at RT. Next, plates were washed thrice with 250 uls/well of TPBS to remove any residual primary antibody. Secondary antibody solution was prepared in 1% non-fat milk in TPBS as well and 100 uls/well was added to the plates for 1 hour at RT. For human samples, anti-human IgG conjugated to horseradish peroxidase (HRP) was used at a dilution of 1:3000 (Millipore Sigma; catalog #A0293). For mouse samples, anti-mouse IgG conjugated to HRP was used at the same dilution (Rockland antibodies and assays; catalog #610-4302). Specifically, a mouse anti-histidine antibody (Takara; catalog #631212) was used as a positive control to detect proteins with a hexa-histidine tag. Once the secondary incubation was done, plates were again washed thrice with 250uls/well of TPBS. Developing solution was made in 0.05M phosphate-citrate buffer at pH 5 using o-phenylenediamine dihydrochloride tablets (Sigma-Aldrich; OPD) at a final concentration of 0.04 mg/ml. One hundred uls/well of developing solution was added to each plate for exactly 10 minutes after which the reaction was halted with addition of 50 uls/well of 3M hydrochloric acid (HCl). Plates were read at an optical density of 490 nanometers using a Synergy 4 (BioTek) plate reader. Eight wells on each plate received no primary antibody (blank wells) and the optical density in those wells was used to assess background. Area under the curve was calculated by deducting the average of blank values plus 3 times standard deviation of the blank values. All data was analyzed in Graphpad Prism 7. This protocol has been described in detail earlier (Bailey et al., 2019; Wohlbold et al., 2015).

Purified monoclonal antibodies were used at a concentration of 30 ug/ml and then subsequently diluted 3-fold. Purified monoclonal antibodies were only incubated on the coated plates for an hour. The remaining part of the protocol was the same as above (Amanat et al., 2020a; Wohlbold et al., 2016).

### Bio-layer Interferometry Binding Experiments

Bio-layer Interferometry (BLI) experiments were performed using the BLItz system (fortéBIO, Pall Corporation). Recombinant human Fc fusion ACE2 (SinoBiological) was immobilized on an anti-human IgG Fc biosensor, and RBDs were then applied to obtain binding affinities. Single-hit concentrations were tested at 5.8 μM for binding. All measurements were repeated in subsequent independent experiments. K_D_ values were obtained through local fit of the curves by applying a 1:1 binding isotherm model using vendor-supplied software. All experiments were performed in PBS pH 7.4 and at room temperature.

### hACE2 competition interferometry experiments

Interferometry experiments were performed using a BLItz instrument (fortéBIO, Sartorius). Polyhistidine-tagged Fabs were immobilized on Ni-NTA biosensors at 10 µg/ml and SARS-CoV-2 RBD was supplied as analyte at 5µM alone or pre-mixed with hACE2-Fc at different concentrations. Maximal signal at association (Rmax) was used to plot the concentration-dependent competition with hACE2. All experiments were performed in PBS at pH 7.4 and at room temperature.

### RBD-hACE2 ELISA

25ng of hACE2-Fc fusion protein expressed in HEK293 cells were adhered to high-capacity binding, 96 well-plates (Corning) overnight in PBS. Plates were blocked with 5% BSA in PBS containing Tween-20 (PBS-T) for 1hr at room temperature (RT). Blocking solution was discarded and 5-fold dilutions of 6xHis-tagged RBDs in PBS were added to wells and incubated for 1hr at RT. Plates were then washed three times with PBS-T. Anti-polyhistidine IgG-Biotin (Abcam) in PBS-T was added to each and incubated for 1hr at RT. Plates were then washed three times with PBS-T. Streptavidin-HRP (Abcam) in PBS-T was added to each and incubated for 1hr at RT. Plates were then washed three times with PBS-T

Plates were developed using 1-Step Ultra TMB substrate (ThermoFisher), stopped with sulfuric acid and immediately read using a plate reader at 450nm. Data were plotted using Prism 9 (GraphPad Software) and affinities determined by applying a nonlinear regression model.

### Viruses and cells

Vero.E6 cells (ATCC #CRL-1586) cells were maintained in culture using Dulbecco’s Modified Eagles Medium (DMEM, Gibco) which was supplemented with 10% fetal bovine serum (FBS, Corning) and antibiotics solution containing 10,000 units/mL of penicillin and 10,000 µg/mL of streptomycin (Pen Strep, Gibco)(10). Wild type SARS-CoV-2 (isolate USA-WA1/2020), hCoV-19/South Africa/KRISP-K005325/2020 (B.1.351, BEI Resources NR-54009) and hCoV-19/England/204820464/2020 (B.1.1.7, BEI Resources NR-54000) were grown in cells for 3 days, the supernatant was clarified by centrifugation at 4,000 g for 5 minutes and aliquots were frozen at -80°C for long term use. The viruses were subjected to deep sequencing to ensure that no mutations had taken place in culture. The polybasic cleavage site changed to WRAR in the B.1.351 variant virus during cultivation in cell culture (as known for this virus at BEI Resources) and no other unexpected mutations occurred. A primary virus isolate, PV14252, bearing mutations and deletions in the spike was obtained by incubating 200 uls of viral transport media from the nasopharyngeal swab with Vero.E6 cells. The sequence of the passage 2 viral isolate was identical to the sequence obtained directly from the clinical specimen. Sequencing was performed on the Illumina platform as described previously (Gonzalez-Reiche et al., 2020). Both replication competent viruses were used to test serum from study participants and antibodies for neutralization activity.

### Neutralization assay

Twenty-thousand cells in 100 uls per well were seeded on sterile 96-well cell culture plates one day prior to the neutralization assay. In general, cells were used at 90% confluency to perform the assay. All serum samples were heat-inactivated to eliminate any complement activity. Serial dilutions of serum samples were made in 1X minimal essential medium (MEM; Life Technologies) starting at a dilution of 1:20. All work with authentic SARS-CoV-2 (isolate USA-WA1/2020 and PV14252) was done in a biosafety level 3 (BSL3) laboratory following institutional biosafety guidelines and has been described in much greater detail earlier (Amanat *et al*., 2020b; Amanat et al., 2020c). Six hundred median cell culture infectious doses (TCID_50_s) of authentic virus (USA-WA1/2020 and PV14252) was added to each serum dilution and virus-serum mixture was incubated together for 1 hour inside the biosafety cabinet. Media from the cells was removed and 120 uls of the virus-serum mixture was added onto the cells for 1 hour at 37°C. After one hour, the virus-serum mixture was removed and 100 uls of each corresponding dilution was added to every well. In addition, 100uls of 1X MEM was also added to every well. Cells were incubated for 48 hours at 37°C after which the media was removed and 150 uls of 10% formaldehyde (Polysciences) was added to inactivate the virus. For assay control, remdesivir was used against both the wild type virus as well as the patient isolate. After 24 hours, cells were permeabilized and stained using an anti-nucleoprotein antibody 1C7 as discussed in detail earlier (Amanat *et al*., 2020b; Sun et al., 2020).

### Cell sorting and flow cytometry

Staining for sorting was performed using cryo-preserved PBMCs in 2% FBS and 2 mM ethylenediaminetetraacetic acid (EDTA) in PBS (P2). Cells were stained for 30 min on ice with CD20-Pacific Blue (2H7, 1:400), Zombie Aqua, CD71-FITC (CY1G4, 1:200), IgD-PerCP-Cy5.5 (IA6-2, 1:200), CD19-PE (HIB19, 1:200), CD38-PE-Cy7 (HIT2, 1:200), and CD3-Alexa 700 (HIT3a, 1:200), all BioLegend. Cells were washed twice, and single plasmablasts (live singlet CD19^+^ CD3^-^ IgD^lo^ CD38^+^ CD20^-^ CD71^+^) were sorted using a FACSAria II into 96-well plates containing 2 µL Lysis Buffer (Clontech) supplemented with 1 U/µL RNase inhibitor (NEB) and immediately frozen on dry ice, or bulk sorted into PBS supplemented with 0.05% BSA and processed for single cell RNAseq.

### Monoclonal antibody (mAb) generation

Antibodies were cloned as described previously (Wrammert et al., 2011). Briefly, VH, Vκ, and Vλ genes were amplified by reverse transcription-PCR and nested PCR reactions from singly sorted plasmablasts using primer combinations specific for IgG, IgM/A, Igκ, and Igλ from previously described primer sets (Smith et al., 2009) and then sequenced. To generate recombinant antibodies, restriction sites were incorporated via PCR with primers to the corresponding heavy and light chain V and J genes. The amplified VH, Vκ, and Vλ genes were cloned into IgG1 and Igκ expression vectors, respectively, as described previously (Nachbagauer et al., 2018; Wrammert et al., 2008). Heavy and light chain plasmids were co-transfected into Expi293F cells (Gibco) for expression, and antibody was purified with protein A agarose (Invitrogen).

### Single-cell RNAseq library preparation and sequencing

Bulk-sorted plasmablasts were processed using the following 10× Genomics kits: Chromium Next GEM Single Cell 5’ Kit v2 (PN-1000263); Library Construction Kit (PN-1000190); Chromium Next GEM Chip K Single Cell Kit (PN-1000286); Chromium Single Cell Human BCR Amplification Kit (PN-1000253), and Dual Index Kit TT Set A (PN-1000215). The cDNAs were prepared after GEM generation and barcoding, followed by GEM RT reaction and bead cleanup steps. Purified cDNA was amplified for 10–14 cycles before cleaning with SPRIselect beads. Then, samples were evaluated on a 4200 TapeStation (Agilent) to determine cDNA concentration. B-cell receptor (BCR) target enrichments were performed on full-length cDNA. Gene expression and enriched BCR libraries were prepared as recommended by the Chromium Next GEM Single Cell 5’ Reagent Kits v2 (Dual Index) user guide, with appropriate modifications to the PCR cycles based on the calculated cDNA concentration. The cDNA libraries were sequenced on Novaseq S4 (Illumina), targeting a median sequencing depth of 50,000 and 5,000 read pairs per cell for gene expression and BCR libraries, respectively.

### Single cell RNAseq analysis

Single-cell RNA sequencing and BCR sequencing data was processed using Cell Ranger v5.0 and the GRCh38-2020 version of the human genome provided by the manufacturer. Total recovered cells by RNA sequencing were V3: 6,608, V5: 5,256, and V6: 6,325 with a mean of 90.64% read mapped to the genome. Count matrices were processed in R (v4.0.2) using the Seurat (v3.2.2) R package (Stuart et al., 2019). Cells were filtered for percentage of mitochondrial genes less than 15% and number features less than 4,000. The three specimen sequencing runs were integrated using log-normalized count values and canonical correlation approach (Stuart *et al*., 2019) with 2,000 variable features. The resulting single-cell object underwent principal component analysis and the top 30 principal components were used for uniform manifold approximation and projection and identifying neighbors. Clustering was performed using a resolution of 0.6. The integrated RNA sequencing object included 12,568 cells with V3: 4,584, V5: 3,523, and V6: 4,461 cells. The filtered contig annotation output of Cell Ranger vdj were loaded into R and processed using the scRepertoire (v1.1.3) R package (Borcherding et al., 2020). Clonotypes were assigned using igraph (v1.2.6) network analysis of components generated from CDR3 sequences greater than or equal to 0.85 normalized Levenshtein distance. Percent of cells expressing genes along the UMAP embedding was visualized using the schex (v1.3.0) R package. For mutation analysis, heavy chains of mAbs and single-cell BCRs first underwent V(D)J gene annotation using IgBLAST (v1.14.0) (Ye et al., 2013) with human reference (release 201931-4) from the international ImMunoGeneTics information system (IMGT) (Giudicelli et al., 2005) and then parsing using Change-O (v0.4.6) (Gupta et al., 2015). Mutation frequency was calculated, as described in (Turner *et al*., 2020), using the “calcObservedMutations” function from SHazaM (v.1.0.2) (Gupta *et al*., 2015) and by counting the number of nucleotide mismatches from the germline sequence in the heavy chain variable segment leading up to the complementary-determining region 3 (CDR3), while excluding the first 18 positions that could be error-prone due to the primers used for generating the mAb sequences.

### Structure visualization and statistical analysis

Structural figures were modeled and rendered in Pymol (The PyMOL Molecular Graphics System, Version 2.4 Schrödinger, LLC). Statistical analysis was performed in GraphPad Prism using a one-way ANOVA with correction for multiple comparisons.

## Figure Legends

**Supplementary Figure 1.**
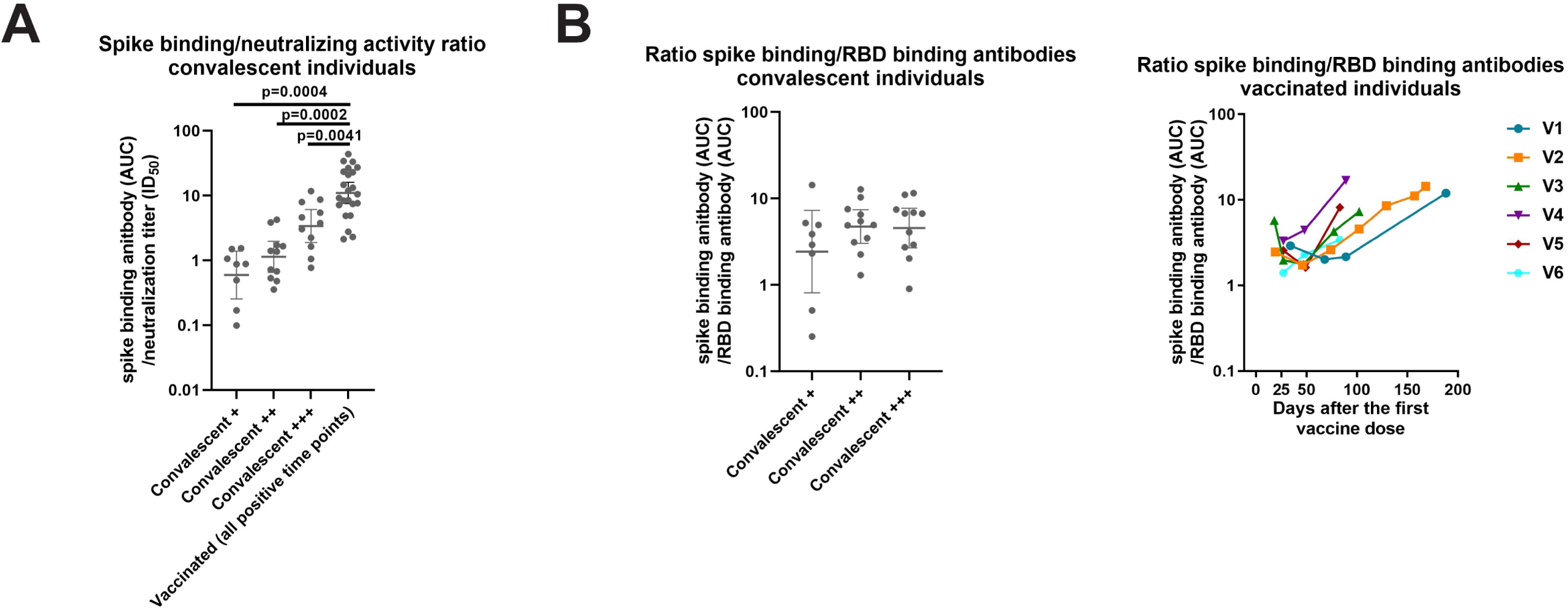
**Full length spike to RBD ratios (A) and comparison of binding to neutralizing titer ratios between naturally infected and vaccinated individuals (B)**.

**Supplementary Figure 2.**
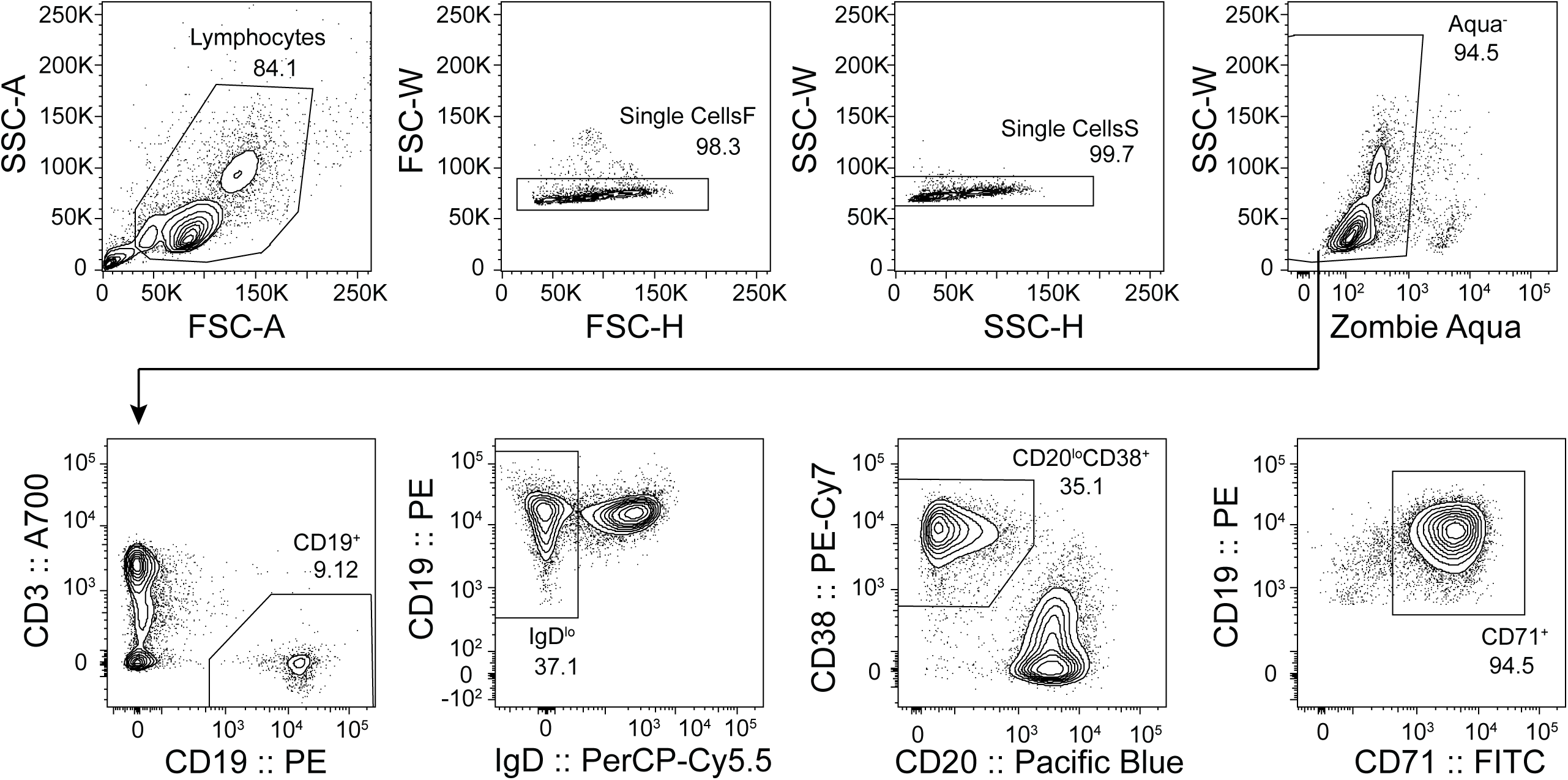
**Gating strategy for sorting plasmablasts from total PBMCs isolated one week after second immunization**.

**Supplementary Figure 3.**
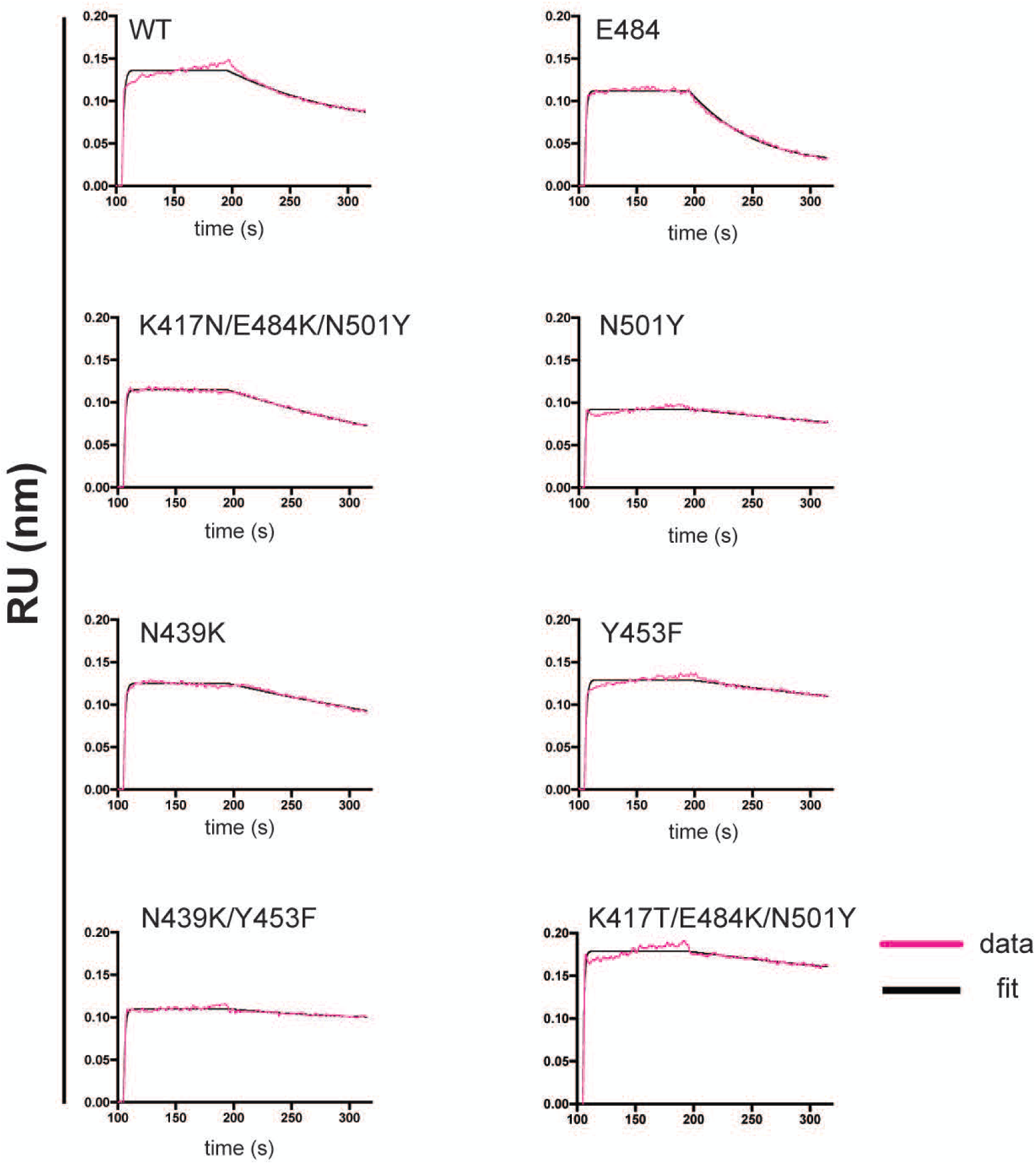
**Representative Biolayer Interferometry binding isotherms from two independent experiments. The raw data are show in pink and the Langmuir 1:1 kinetics fit is show in black**.

**Supplementary Figure 4.**
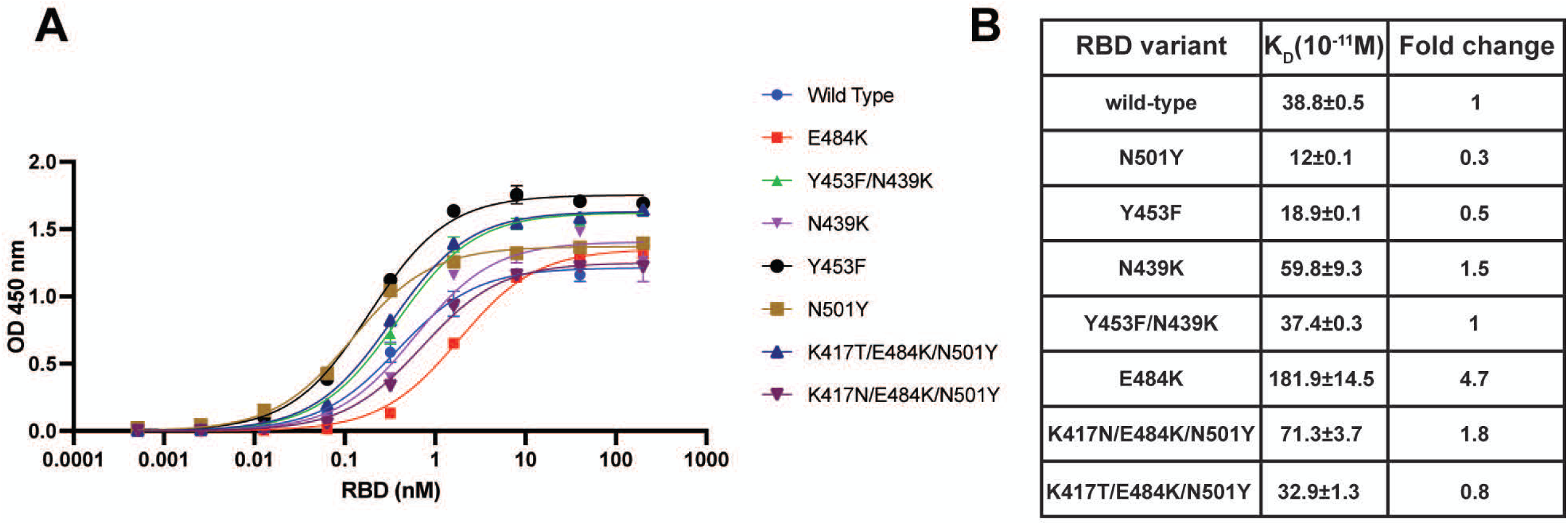
Binding of SARS-CoV-2 variant RBDs to ACE2. A. ELISA curves of the RBD variants binding to human ACE2. Shown are the binding curves calculated with nonlinear regression to the arithmetic mean values from eight replicates ± SEM. The calculated steady-state K_D_ values ± SEM from end-point ELISA measurements and the fold-change in comparison to wild type RBD are reported in **B**.

**Supplemental Table 1:** Study participant and biospecimen information.

| Vaccinees | Spike IgG response<sup>1*</sup> | Sex | Age group (yrs) | Specimen tested |
| --- | --- | --- | --- | --- |
| V1 | Strong positive | F | >60 | several longitudinal time points including days 34, 68, 89 and 188 post vaccination |
| V2 | Strong positive | M | 30-40 | several longitudinal time points including days 19, 47, 74, 102, 129, 157 and 186 post vaccination |
| V3 | Strong positive | F | 50-60 | several longitudinal time points including days 19, 27, 47, 77 and 102 post vaccination |
| V4 | Strong positive | M | >60 | several longitudinal time points including days 27, 48 and 89 post vaccination |
| V5 | Strong positive | F | 40-50 | several longitudinal time points including 27, 49 and 83 days post vaccination |
| V6 | Strong positive | F | 30-40 | several longitudinal time points including 27, 48 and 83 days post vaccination |
| **Seronegative, post pandemic** |  | **Sex** | **Age group (yrs)** | **Days from last negative serology test when the sample was taken** |
| N1 | Negative | F | 40-50 | 23 |
| N2 | Negative | F | 20-29 | 24 |
| N3 | Negative | F | 20-29 | 23 |
| N4 | Negative | F | 30-35 | 22 |
| **Seropositive, natural infection** |  | **Sex** | **Age group (yrs)** | **Days post onset of COVID-19 symptoms when the sample was taken** |
| P1 | Weak positive | M | 20-29 | 260 |
| P2 | Weak positive | M | 50-59 | no data available |
| P3 | Weak positive | F | 30-39 | 111 |
| P4 | Weak positive | F | 30-39 | 221 |
| P5 | Weak positive | F | 30-39 | 254 |
| P6 | Weak positive | F | 20-29 | 247 |
| P7 | Weak positive | M | 30-39 | 220 |
| P8 | Weak positive | F | 20-29 | Asymptomatic |
| P9 | Moderate positive | M | 30-39 | no data available |
| P10 | Moderate positive | F | 30-39 | 197 |
| P11 | Moderate positive | F | 50-59 | Asymptomatic |
| P12 | Moderate positive | F | 30-39 | Asymptomatic |
| P13 | Moderate positive | M | 30-39 | 234 |
| P14 | Moderate positive | F | 20-29 | 273 |
| P15 | Moderate positive | M | 30-39 | Asymptomatic |
| P16 | Moderate positive | F | 20-29 | 258 |
| P17 | Moderate positive | F | 20-29 | 246 |
| P18 | Moderate positive | M | 20-29 | Asymptomatic |
| P19 | Moderate positive | F | 50-59 | 204 |
| P20 | Strong positive | F | 50-59 | no data available |
| P21 | Strong positive | F | 30-39 | 245 |
| P22 | Strong positive | M | NA | 170 |
| P23 | Strong positive | F | >60 | Asymptomatic |
| P24 | Strong positive | F | 40-49 | no data available |
| P25 | Strong positive | F | 50-59 | 191 |
| P26 | Strong positive | F | 30-39 | no data available |
| P27 | Strong positive | F | 50-59 | 113 |
| P28 | Strong positive | M | >60 | Asymptomatic |
| P29 | Strong positive | M | 18-19 | 218 |
| P30 | Strong positive | M | 50-59 | 219 |

<sup>1</sup>Samples were categorized based on initial titers obtained from Mount Sinai's CLIA laboratory test. Weak positive: 1:80 – 1:160 weak positive; 1:320-1:960 moderate positive; 1:960-1: ≥2880 strong positive
\* All six vaccinees were sero-negative for SARS-CoV-2 without clinical evidence of COVID10 prior to SARS- CoV-2 spike mRNA vaccination.

**Supplemental Table 2:**
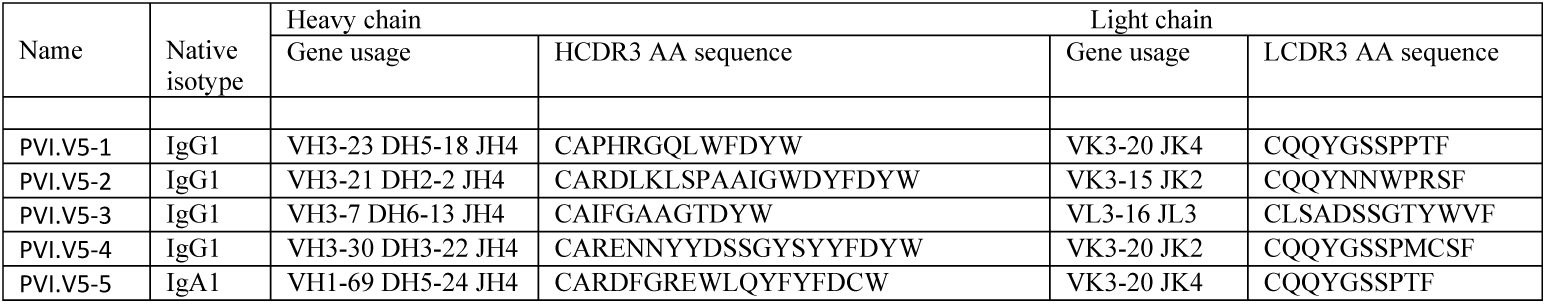
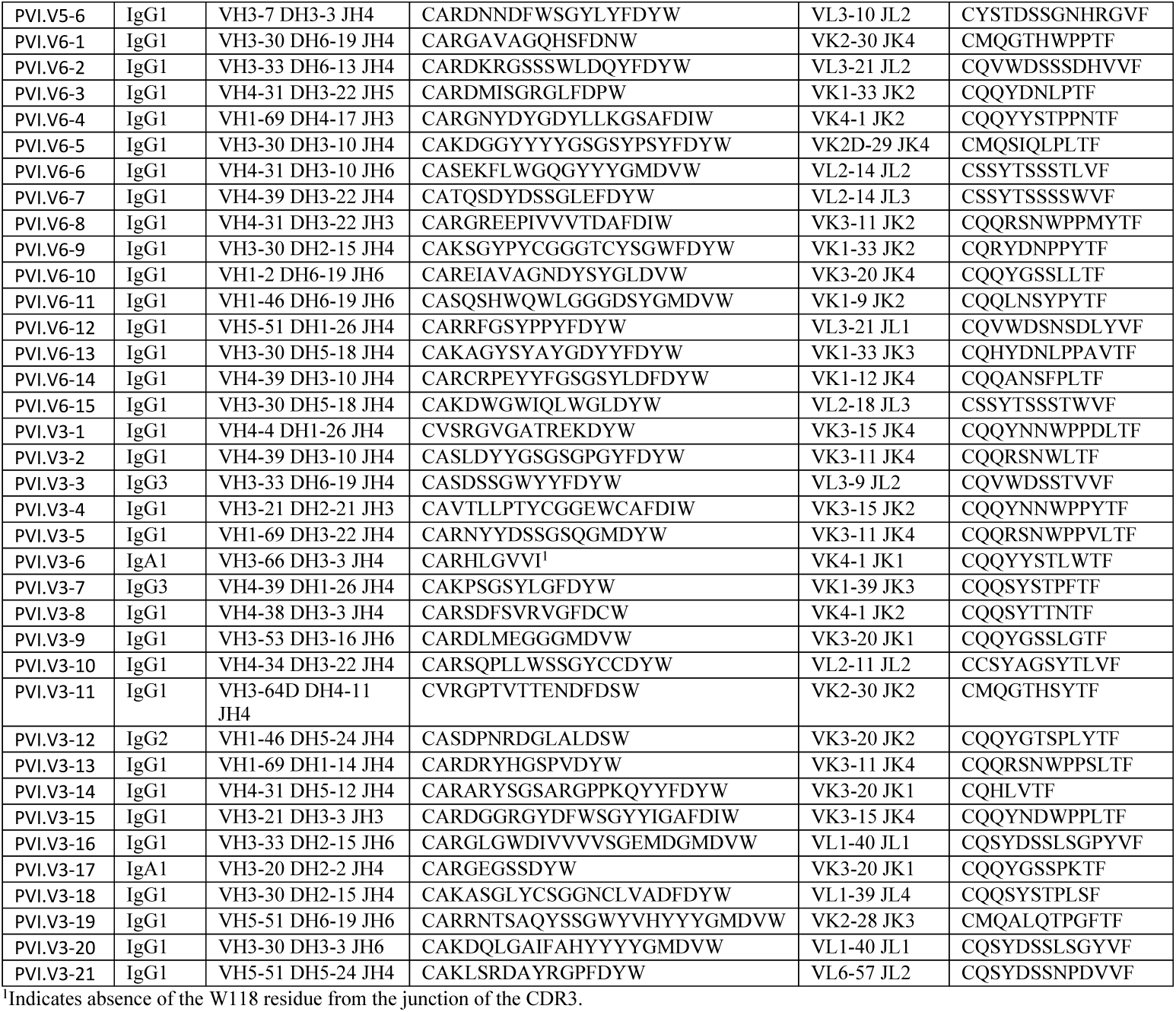
Immunoglobulin gene usage of the spike-mAbs.

